# Predicting Self-harm Incidents Following Inpatient Visits using Disease Comorbidity Network

**DOI:** 10.1101/2020.02.03.20019331

**Authors:** Zhongzhi Xu, Qingpeng Zhang, Paul Siu Fai Yip

**Affiliations:** School of Data Science, City University of Hong Kong, Hong Kong, China; Centre for Suicide Research and Prevention, The University of Hong Kong, Hong Kong S.A.R., China

## Abstract

Self-harm is serious but preventable, particularly if the risk can be identified early. But the early detection of self-harm individuals is so far not satisfied. This study aims to develop and test a comorbidity network-enhanced deep learning framework to improve the prediction of individual self-harm within 12 months after hospital discharge. Between January 1, 2007, and December 31, 2010, we obtained 2,323 patients with self-harm clinical record from 1,764,094 inpatients across 44 public hospitals in Hong Kong and 46,460 randomly sampled population controls from those same hospitals. Eighty percent (80%) of the study sample was randomly selected for model training, and the remaining 20% was set aside for model testing. The proposed comorbidity network-enhanced model was compared with a baseline deep learning model for self-harm prediction. The C-statistic, precision and sensitivity were used to evaluate the prediction accuracy of the proposed model and the baseline model. Experiments demonstrated that the proposed comorbidity network-enhanced model outperformed baseline model in identifying patients who would self-harm within 12 months (C-statistic of proposed model 0.89). The precision was 0.54 for positive cases and 0.98 for negative cases, whilst the sensitivity was 0.72 for positive cases and 0.96 for negative cases. Results indicated that it is critical to consider the general disease comorbidity patterns in self-harm screening and prevention programs. The model also extracted the most predictive diagnoses, and pairs of comorbid diagnoses which provide medical professionals with an effective screening strategy.

## Introduction

Self-harm, which includes non-fatal intentional self-poisoning/overdose and self-injury, is a significant public health issue in many countries.^1–5^ It usually leads to tragic outcomes, and unless detected early and averted, is linked to risk of future suicide.^6, 7^ Self-harm and its sequelae can place significant burdens on healthcare and societal resources. Thus, the early detection of people at risk of self-harm is important for medical and social health professionals.^8,9^

Currently, attempts to prevent self-harm have relied mostly on self-reporting of self-harm thoughts and intentions.^10,11^ This may not capture the extent of the problem, because self-reported data are subject to reporting biases. This is particularly so for reporting self-harm attempts, because of people’s fear of potential prejudice and discrimination, and their desire to avoid hospitalization and public scrutiny.^12^

The notion of mining diagnostic information in electronic health records for self-harm and suicidal behaviour prediction has been recently explored.^12,13,14^ The rationale for using clinical diagnoses for the prediction of self-harm is that future self-harm events, and patients’ historical medical diagnoses, have been found to be associated. Such associations may come from common underlying biological dysfunctions and/or persisting social factors. For example, Tran et al.^13^ identified that historical medical diagnoses (coded in ICD-10) Asphyxiation (T71), Organic amnesic syndrome (F04) and Very high alcohol level in blood (Y90.5-Y90.8, Y91.3) are correlated with the risk for self-harm and suicidal behaviour. In another study,^15^ patients diagnosed with cancer have 1.51 times higher risk for self-harm than fully matched general population.

Despite understandings of disease consequences, there is limited capacity to reliably predict self-harming behaviours. This is because risk prediction models generally concentrate on the main effects of medical diagnoses to determine self-harm risk. However, recent research has found that the co-occurrence of multiple diseases is related to self-harm risk in a different way from simply adding the main effects of individual diseases. For example, diseases of the nervous system and past self-harm attempts interact strongly when quantifying risk of self-harm, because diseases of the nervous system may exaggerate the effect of past self-harm behaviour^16^. As a result, the self-harm risk profile of a patient who has both past self-harm attempts, and a nervous system disease, is much higher than the additive effects of the separate risk estimates.^16^ Traditional regression models can introduce pairwise interactions to address this issue. However, this approach remains limited because (a) the pairwise interactions are usually difficult to effectively predefine, particularly when there are many correlates; and (b) the complex interactions between characteristics of multiple diseases may go beyond pairwise interactions.

Moreover, disease comorbidity patterns may not be well captured by using traditional disease risk prediction frameworks because the set of diagnoses and their interactions to evaluate is large and complex and the sample size of self-harm studies is usually small. This will result in the learning of model parameters not smooth and the capture of interacting patterns difficult.

To address the complex, potentially multiplicative effects of diagnoses on self-harm risk, we proposed the use of existing side information - disease comorbidity networks, to enhance the effectiveness of risk prediction for self-harm. The comorbidity network refers to existing knowledge of the frequency of the co-occurrence of diseases. The introduction of the comorbidity network presents an exciting opportunity to improve the accuracy of self-harm prediction by synthesizing knowledge from various sources of information. So far, this resource is becoming increasingly available worldwide but has been surprisingly under-utilised by healthcare researchers.

## Methods

### Study population

The study examples came from electronic healthcare records (EHR) collected from Hong Kong residents, admitted to the 44 public hospitals in Hong Kong between January 2007 and December 2010. This historical dataset was chosen because it incurred minimum ethical risk of identification to individuals whose data was captured within it. A unique patient identifier, gender, age, disease codes of diagnoses, date of admission and date of hospital discharge were available for each inpatient admission. This dataset contained over 5.2 million electronic health records (visits) covering 1,764,094 inpatients. The disease codes were defined by the International Classification of Diseases, Ninth Revision, Clinical Modification (ICD-9-CM). The data was anonymized.

Diagnoses of self-harm were identified using ICD-9CM codes E950–E959 (suicide and self-inflicted injury).^16^ We aimed to identify patients with high self-harm risk in the year subsequent to their last clinical diagnoses. Therefore, the self-harm samples included any patient with a self-harm diagnosis and at least one admission recorded in the previous twelve months. Patients younger than 10 years were excluded. From the larger dataset, 46,460 patients without self-harm records were randomly selected as the counterparts, making the ratio between positive (self-harm) and negative (no self-harm) cases 1:20.^17^ Finally, 2,323 self-harm samples and 46,460 counterparts were extracted as study samples. Eighty percent (80%) of the study sample was randomly selected for model training, and the remaining 20% was set aside for model testing. Figure 1 shows the data sampling flow. Age, gender and patients’ historical clinical diagnoses were used as the predictors for self-harm and suicidal behaviours.

**Figure 1.**
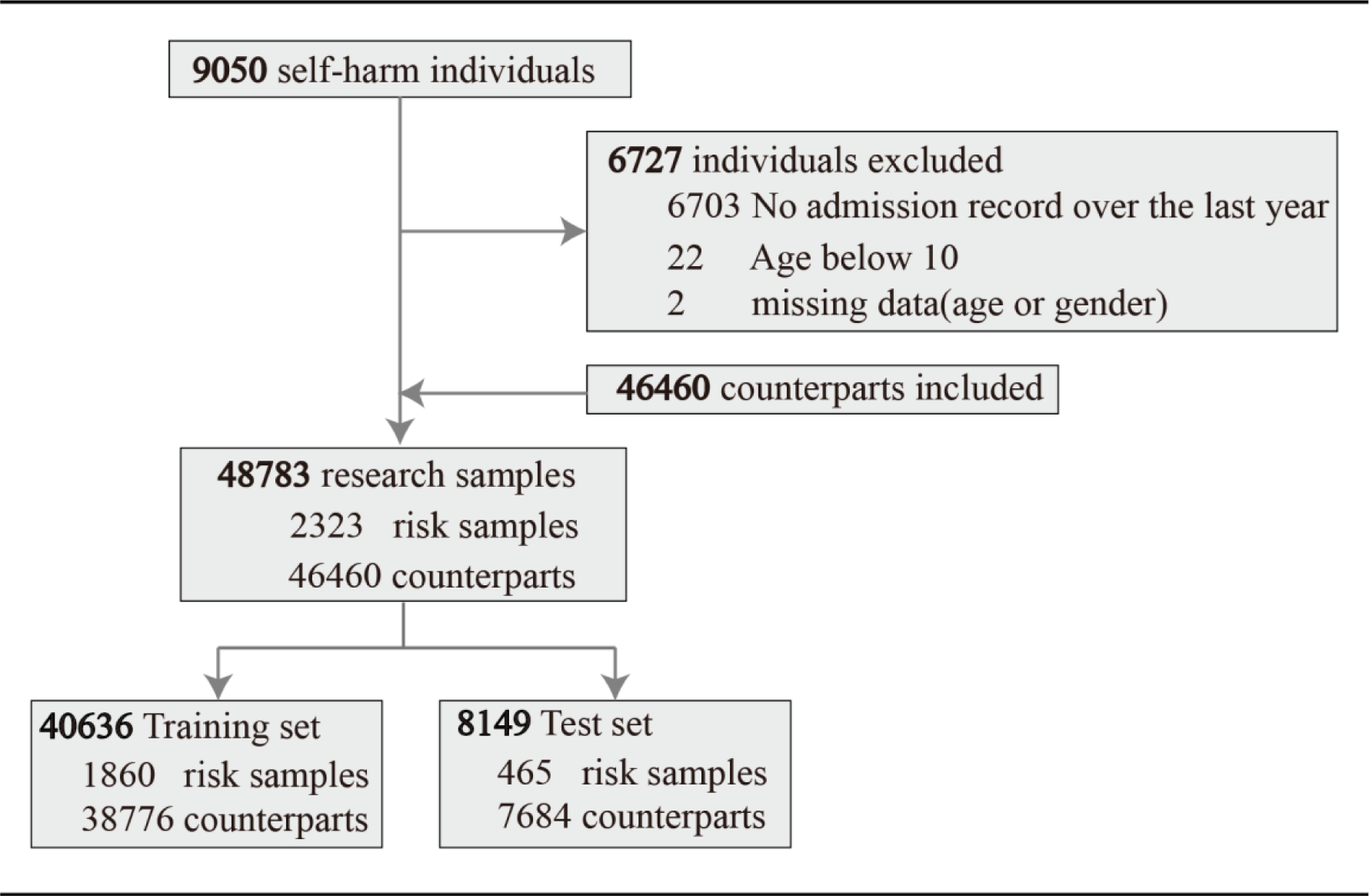
Flowchart of selecting research examples.

To ensure sufficient numbers for analysis, we trimmed the five-digit ICD-9-CM code to three digits. Sparse data leads to significantly higher computational cost and over-fitting issues. Both this study and previous studies show that three digits are sufficiently informative to discriminate patients from each other in downstream tasks.^18,19^

### Patient and Public Involvement

Because this study was secondary EHR data analytics, it was not appropriate or possible to involve patients or the public in the design, or conduct, or reporting, or dissemination of our research. The data is provided by the Hospital Authority of Hong Kong the ethical approval UW11-495. The data can not be made available to others according to the Hospital Authority and the ethical approval.

### Disease comorbidity networks

Comorbidity relationships can be defined using different scales, such as co-occurring frequency, shared expressed genes, shared disease proteins, shared single-nucleotide polymorphisms, and shared pathways involved in both diseases^20^. In this study, we defined the existence of a comorbidity relationship between two diseases as the frequency of co-occurrence in an inpatient visit.^21^ This intuitive approach is more appropriate for this research because self-harm is associated with many diseases, leading to an elevated risk for self-harm even if the diseases do not share physiological relations. For example, people may engage in self-harm because of the desperate feeling brought by cancer.^22^ In addition, much of the physiological relations between diseases remain unexplored.^23^ Thus we explored the co-occurrence relations to have a comprehensive understanding of the comorbidity patterns that lead to self-harm.^21^

Hong Kong public hospital EHR can record up to 15 disease diagnoses for each inpatient admission. The average number of diagnoses for each admission is 2.27 (standard deviation (SD) 0.51). We defined the comorbidity network as an undirected graph, with each node representing a disease diagnosis. The weighted edge between two nodes represented the co-occurrence frequency of the two corresponding diseases. The comorbidity network was constructed using 7-year EHR data (January 2000 to December 2006). There were 938 nodes (disease diagnoses) and 15,846 weighted edges (co-occurring relations). This comorbidity network represented empirical evidence of the disease interactions in the population in Hong Kong during the study period. To the best of our knowledge, this is the most comprehensive comorbidity network based on real-world EHR data. We believe that this comorbidity network is generic and can be applied to other clinical decision-making problems. A visualization of the network is provided in Figure 2. Colours of nodes represent 19 disease categorizations according to the ICD-9CM. This network can then be incorporated into downstream self-harm prediction task by representing each disease as a low-dimensional vector. Refer to the following section for detailed information.

**Figure 2.**
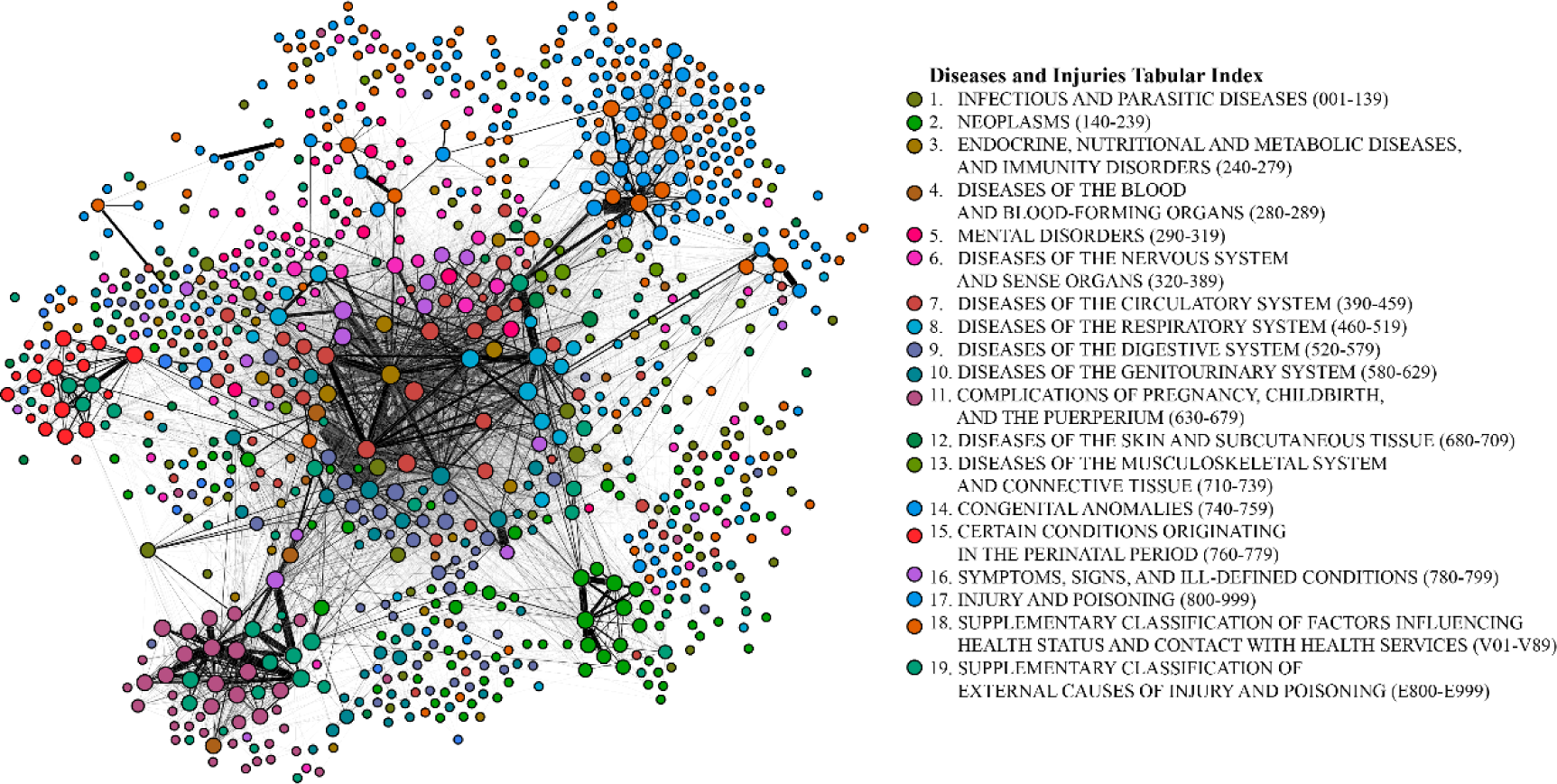
Comorbidity disease network. Edges weighted less than 10 are hidden for simplicity.

### Model

This section introduces a novel patient embedding method, namely the *Dx2vec* (“*diagnoses to vector*”), to represent simultaneously the diagnoses, the comorbidity patterns among diagnoses, and the temporal patterns of historical inpatient admissions for each patient as a low-dimensional feature vector. The Dx2vec is the first comprehensive embedding method that considers these three pieces of critical information for downstream applications. Dx2vec translate such multi-aspect information about a patient into a low-dimensional vector while still preserving the critical information hidden in patients’ historical EHR, so that it enables higher-resolution modeling for predicting self-harm.

There are four steps to generate the Dx2vec embedding (Figure 3). First, we adopted a classic network embedding method, DeepWalk,^24^ to learn the feature vectors for each disease (namely, the *disease embedding* (blue box in Figure 3)) from the comorbidity network. Specifically, for each disease in the network, we assigned fifty random walks from the corresponding node and calculate the likelihood of reaching other nodes (i.e. closeness between diseases). Those likelihood values were used to learn the disease’s embedded feature vector, which in turn preserved the relationships between diseases. Each dimension in the feature vector represented a latent feature of the disease. These latent features captured the comorbidity patterns not explicitly represented by the existing dataset (such as lifestyle and social status). Interpreting each latent feature was possible but required much more information, which was often unavailable in practice. Figure 4A provides an example of learned disease embeddings of three categories of diseases in a three-dimensional space. It is obvious that diseases in the same category are clustered into the same group in this three-dimensional space. This demonstrates that the learned disease embedding encodes the relations among diseases in the comorbidity network. Instead of using indicators where diseases are represented independently, we use disease embedding-vectors that preserve the information of the comorbidity network. We hypothesized this would contribute to the increase of model prediction power.

**Figure 3.**
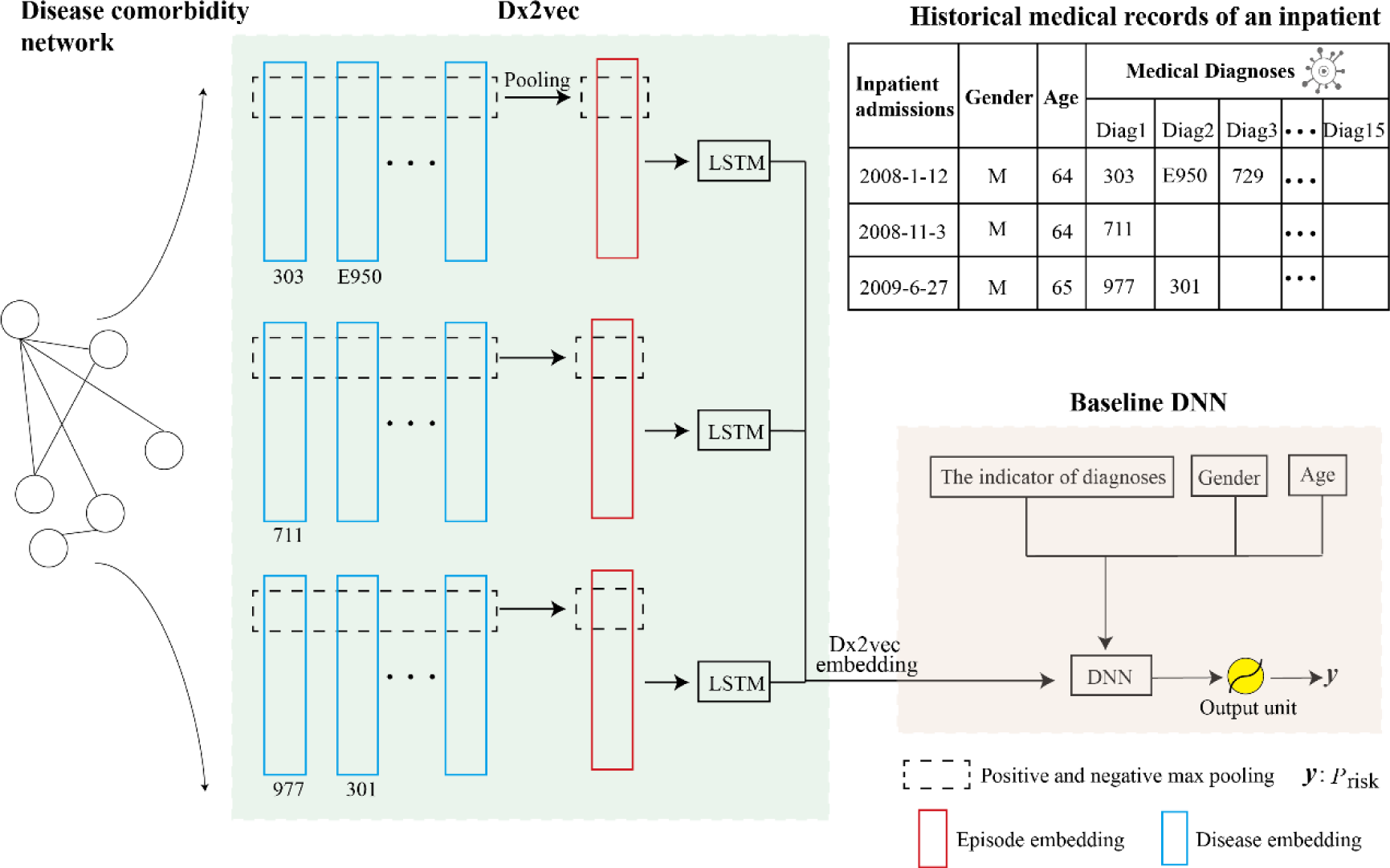
The architecture of the Dx2vec-based DNN model and the baseline DNN model.

**Figure 4.**
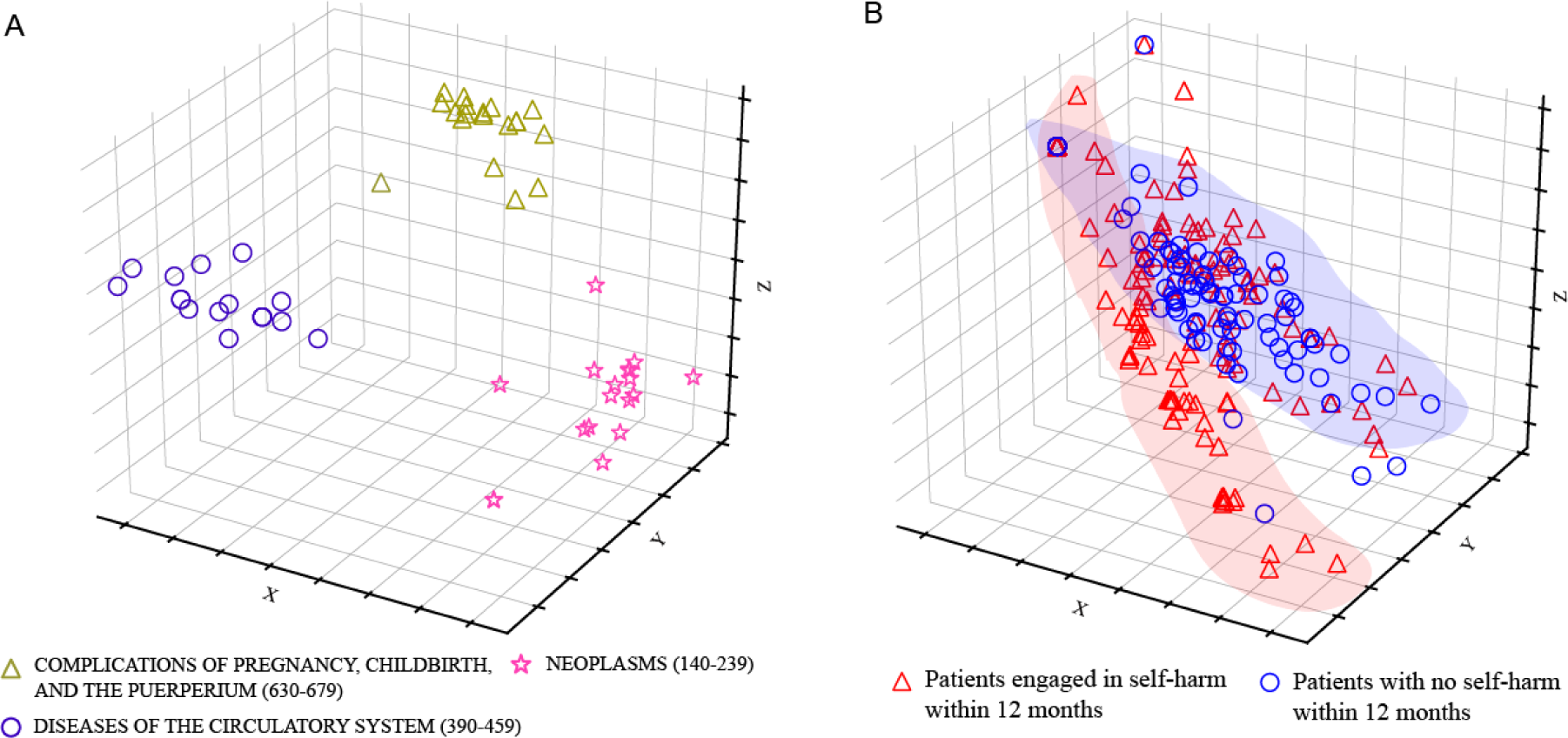
*A*. Disease embedding of three categories of diseases in a three-dimensional space. *B*. The Dx2vec embedding of 200 patients in a three-dimensional space. 100 patients would self-harm within 12 months (red triangles), and 100 would not (blue circles)

Secondly, we generated the embedding for each inpatient admission episode (namely, the *episode embedding* (red box in Figure 3)) through positive-and-negative max pooling of the disease embedding for all diagnoses in this episode. More specifically, for each dimension (black dash box in Figure 3), we set the value to be the one with the largest absolute value among all in this dimension for all diagnoses. For example, for a patient with two diagnoses *A* and *B*, which is represented using a 2-dimensional disease embedding ***v***_*A*_ = (1.2, 0.2)^T^ and ***v***_*B*_ = (−1.5, 0.1)^T^, respectively. The positive-and-negative max pooling is then (−1.5, 0.2)^T^. In this way, the obtained episode embedding captured the most distinct characteristic in each feature dimension.

Thirdly, episode embedding vectors were fed into a deep learning module, more specifically, the LSTM (Long Short-Term Memory) unit, to learn the final Dx2vec embedding (output of the green dash box module in Figure 3). The LSTM module captured the temporal patterns of multiple inpatient admission episodes and the associated disease comorbidity. As the output of the LSTM module, Dx2vec embedding is an innovative way to represent the multiplicity of diagnoses, comorbidity patterns among diagnoses, and the temporal patterns of historical inpatient admissions for each patient. Given the low dimensionality of Dx2vec embedding, it was thus feasible to incorporate these complex patterns into downstream deep learning tasks.

The majority (91%) of the patients in the dataset had three or fewer historical inpatient admission episodes. Moreover, remote episodes may introduce noisy information that might not relevant to self-harm occurring in the near future. Therefore, we only modelled the most recent three admission records into the model.

Figure 4B presents a three-dimensional Dx2vec embedding of 200 randomly sampled cases and counterparts (100 patients would self-harm within 12 months, 100 would not). This figure demonstrates that the three-dimensional Dx2vec embedding can roughly distinguish these two groups of patients. Note in this figure that there are still many red and blue cases mixed together. We can use higher dimensional Dx2vec embedding to further distinguish them. In the main model to be implemented, the Dx2vec embedding has 64 dimensions.

The Dx2vec embedded vector is concatenated with the indicators of diagnoses, age, and gender of the patients. The final concatenated vector is then fed into a deep neural network (DNN) to generate the final risk prediction. We also adopt a traditional DNN model without Dx2vec embedding (dash brown box module in Figure 3) as the baseline.

## Results

### Predicting the risk of self-harm

To develop a comprehensive understanding of the model performance in the imbalanced dataset, it was necessary to consider multiple metrics from different perspectives. We report the precision, sensitivity and C-statistic metrics in this paper. We defined the patients with no self-harm attempt within 12 months as negative cases, and the patients who engaged in self-harm within 12 months as positive cases. Unsurprisingly, both Dx2vec-based DNN model and the baseline DNN model achieved a high precision of the negative class 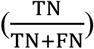 and high sensitivity of the negative class 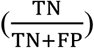 due to the imbalanced nature between positive and negative examples. For positive cases, which are really what we care for in practice, the precision 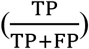 of Dx2vec-based DNN model and baseline DNN model was similar (0.54 vs 0.53, shown in Figure 5A and C), indicating that there was greater than 50% chance that these deep learning models’ predictions were correct. Given the imbalanced nature of the data and the extreme consequences of positive cases, some false alarms were reasonable and acceptable. More importantly, we found that the Dx2vec-based DNN model can significantly improve the sensitivity of positive class 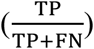 over the baseline DNN (0.72 vs 0.65, also shown in Figure 5A and C). This indicates that the proposed Dx2vec-based DNN model identified more patients with the risk for future self-harm through incorporating the comorbidity patterns of diseases hidden in the comorbidity network. This is critical to real-world clinical practice, to avoid missing patients who are at risk for severe outcomes.

**Figure 5.**
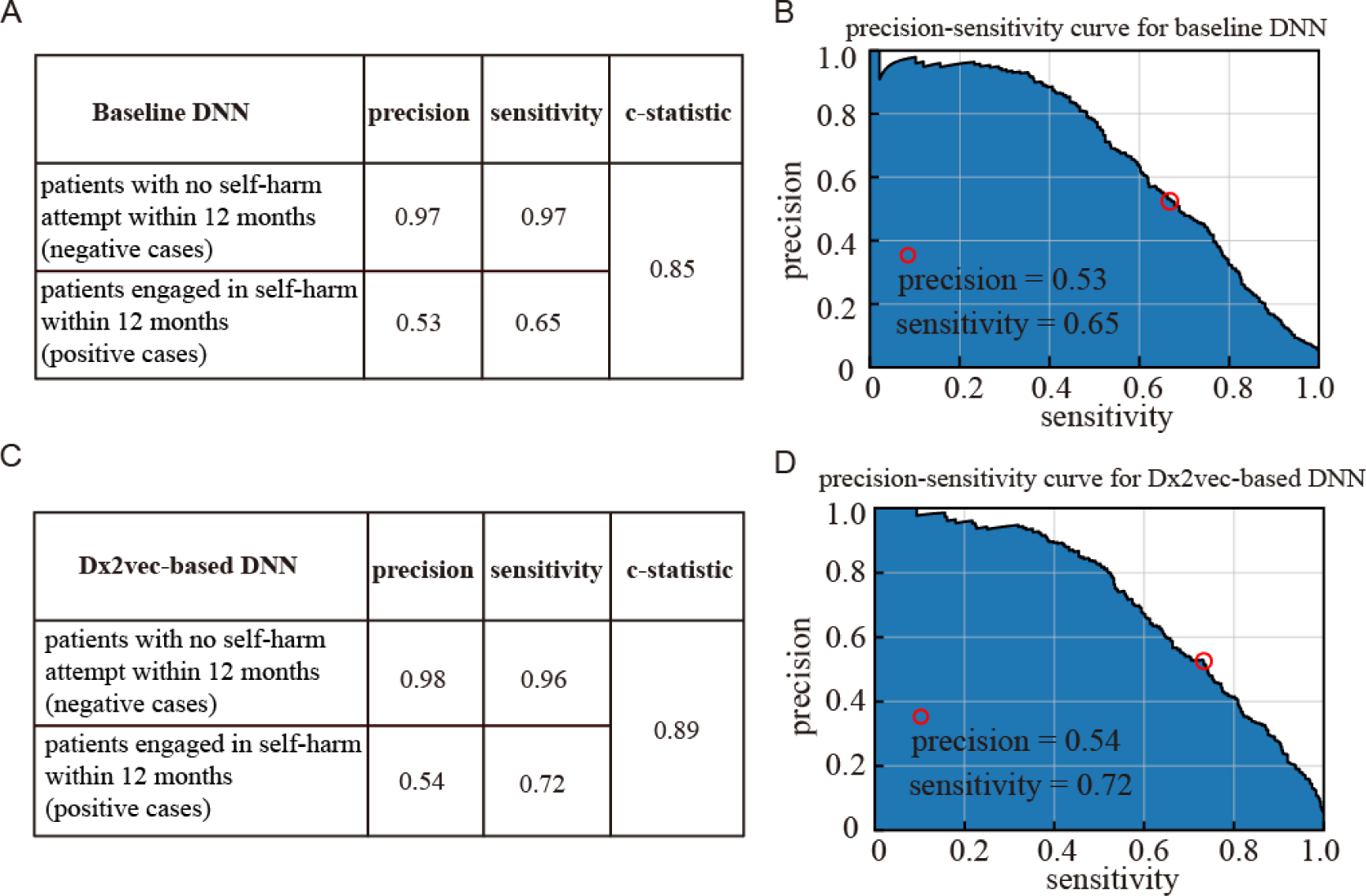
Prediction performance. *A* and *C*. The precision and sensitivity of the positive and negative cases of the Dx2vec-based DNN model and baseline DNN model; *B* and *D*. The precision-sensitivity curves of the positive cases of the two models.

The C-statistics for the Dx2vec-based model and baseline DNN model were 0.89 and 0.85 respectively, again showing the advantage of incorporating the comorbidity network. The precision-sensitivity curves of the positive class for both models are also shown in Figure 5B and D to illustrate the variance of sensitivity and precision.

### Crucial risk factors leading to high self-harm risk

To further understand the prominent risk factors of self-harm, we calculated the predicted probabilities corresponding to each diagnosis/diagnosis pairs by feeding all possible diagnoses/diagnosis pairs into the proposed model iteratively, and obtaining the most predictive diagnoses and pairs of diagnoses in Table 1 and Table 2. These high-risk single and pairwise diagnoses provide straightforward information for healthcare professionals to understand the risk for individual patients, and thus could serve as the basis for interventions to prevent future self-harm.

**Table 1.** Crucial risk factors leading to high self-harm risk.

| Rank | Predictive diagnoses |
| --- | --- |
| 1 | E950 Suicide and self-inflicted poisoning by solid or liquid substances |
| 2 | 989 Toxic effect of other substances, chiefly nonmedicinal as to source |
| 3 | 967 Poisoning by sedatives and hypnotics |
| 4 | E852 Accidental poisoning by other sedatives and hypnotics |
| 5 | E958 Suicide and self-inflicted injury by other and unspecified means |
| 6 | 965 Poisoning by analgesics, antipyretics, and antirheumatics |
| 7 | E952 Suicide and self-inflicted poisoning by other gases and vapours |
| 8 | 292 Drug-induced mental disorders |
| 9 | 977 Poisoning by other and unspecified drugs and medicinal substances |
| 10 | 305 Nondependent abuse of drugs |
| 11 | 986 Toxic effect of carbon monoxide |
| 12 | 303 Alcohol dependence syndrome |
| 13 | 969 Poisoning by psychotropic agents |
| 14 | E866 Accidental poisoning by other and unspecified solid and liquid substances |
| 15 | E868 Accidental poisoning by other utility gas and other carbon monoxide |
| 16 | 304 Drug dependence |
| 17 | 308 Acute reaction to stress |
| 18 | E860 Accidental poisoning by alcohol |
| 19 | E934 Agents primarily affecting blood constituents |
| 20 | 301 Personality disorders |

**Table 2.**
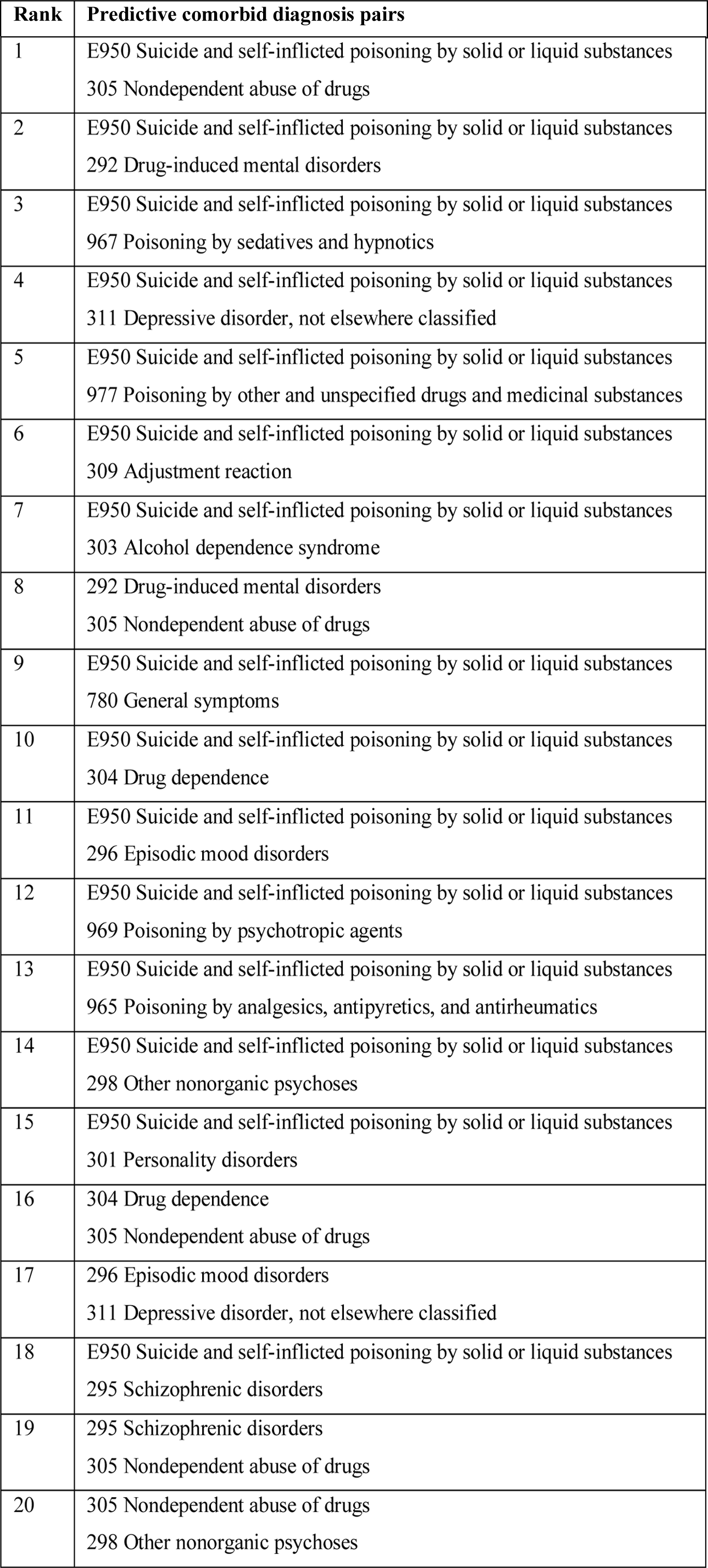
Crucial comorbid diagnosis pairs leading to high self-harm risk

## Discussion

Using data commonly available in EHR, the proposed Dx2vec-based DNN model were able to identify 72% of all inpatients at risk for self-harm within 12 months, with 54% precision. The increasingly widespread adoption of EHR provides unprecedented opportunities for practical application of precision medicine, including the possibility of risk prediction for major health outcomes such as self-harm and suicide death.

Our comorbidity network-aware framework has a number of key strengths. First, it successfully incorporated the disease comorbidity network to enhance the predicting performance. Second, it enabled higher-resolution modeling through considering simultaneously the diagnoses, the comorbidity patterns among diagnoses, and the temporal patterns of historical inpatient admissions for each patient. Third, the proposed disease comorbidity network is generic and can be applied to other models. Last but not least, the proposed framework is also generic and has high flexibility in predicting other health outcomes.

We also looked into details the historical diagnoses associated with future self-harm. They can be clustered into three distinct categories: (a) mental disorders, including 292, 301, 303, 304 and 305; (b) previous self-harm behaviours, including E950, E952 and E958; and (c) diagnoses that might be under-diagnosed, including 965, 967, 969, 977, 986, 989, E852, E860, E866, E868 and E934. These prominent risk factors of self-harm for Hong Kong patients are similar to previous studies in other countries/regions.^10,25,26^ Moreover, among these risk factors identified in our study, we observed some diagnoses, such as 986 (Toxic effect of carbon monoxide), that have rarely been reported in previous studies. A possible explanation is that this diagnosis is resulted by charcoal-burning suicide attempts, which is more popular in Asian countries, especially in Hong Kong.^27^

Moreover, besides diagnoses that explicitly indicated past self-harm attempts (category (b)), many underdiagnoses (category (c)) were also detected to be crucial risk factors. A possible explanation is that people may deny self-harm ideation in order to avoid special treatment and potential prejudice.^28^ Stigmatization medical and health workers against people who self-harm is not uncommon. Understanding of under-diagnosed self-harm phenotypes is important to the identification of self-harm cases.

Our model can provide clinicians with not only the independent effect of individual diagnoses on self-harm, but also the combined effect of multiple diagnoses. Such effects are obtained from comorbidity patterns and are often different from simply adding the independent effects. These identified joint-effects (Table 1) provide data-driven insights on how the risk is elevated if a patient has multiple diagnoses. Most of the predictive comorbid diagnosis pairs involve previous self-harm record (E950). Besides, our model found that if a patient had both 292 (Drug-induced mental disorders) and 305 (Nondependent abuse of drugs), his or her risk for self-harm was even higher than many patients with previous self-harm record (E950). Substance abuse was a major concern especially among young people for the period of study. There are many such observations (in supplementary information Tables S1 and S2), which present helpful insights for clinicians and researchers to design new randomized controlled trials regarding self-harm prevention. More precise prediction and interpretation of the comorbidity patterns can inform more rational and accurate clinical decision support.

Our study has limitations. Specifically, the model was trained on inpatient data that contained only basic demographic information and medical diagnoses. Although the model performance was encouraging, there is additional information (outpatient records, medications, procedures, family histories, genetic factors, prior major life milestones, and social support) about self-harm which could improve its predictive capacity. Now that the generic model procedures have been developed and tested, incorporating addition risk factor information is the next step in model refinement.

## Data Availability

The data is provided by the Hospital Authority of Hong Kong the ethical approval UW11-495. The data can not be made available to others according to the Hospital Authority and the ethical approval.

## Contributors

ZX and QZ formulated the idea. ZX performed the literature review. ZX and QZ developed the model and conducted the experiments. ZX, QZ and SFPY analysed and interpreted the results. ZX, QZ and SFPY wrote the article. All authors had full access to all data (including statistical reports and tables) in the study and take responsibility for the integrity of the data and the accuracy of the data analysis.

